# The relationship between pathological brain activity and functional network connectivity in glioma patients

**DOI:** 10.1101/2023.04.20.23288814

**Authors:** Mona LM Zimmermann, Lucas C Breedt, Eduarda GZ Centeno, Jaap C Reijneveld, Fernando AN Santos, Cornelis J Stam, Marike R van Lingen, Menno M Schoonheim, Arjan Hillebrand, Linda Douw

## Abstract

**Background:** Glioma is associated with pathologically high peritumoral brain activity, which relates to faster progression. Functional connectivity is disturbed locally and throughout the entire brain, associating with symptomatology. We, therefore, investigated how local activity and network measures relate to better understand how the intricate relationship between the tumor and the rest of the brain may impact disease and symptom progression.

**Methods:** We obtained magnetoencephalography in 84 *de novo* glioma patients and 61 matched healthy controls. The offset of the power spectrum, a proxy of neuronal activity, was calculated for 210 cortical regions. We calculated patients’ regional deviations in delta, theta and lower alpha network connectivity as compared to controls, using two network measures: clustering coefficient, a measure of local connectivity, and eigenvector centrality (integrative connectivity). We then tested group differences in activity and connectivity between peritumoral, contralateral homologue regions, and the rest of the brain. We also correlated regional offset to connectivity.

**Results:** As expected, patients’ peritumoral activity was pathologically high, and patients showed higher clustering and lower centrality than controls. At the group-level, regionally high activity related to high clustering in controls and patients alike. However, within-patient analyses revealed negative associations between regional deviations in brain activity and clustering, such that pathologically high activity coincided with low network clustering, while regions with ‘normal’ activity levels showed high network clustering.

**Conclusions:** Our results indicate that pathological activity and connectivity co-localize in a complex manner in glioma. This insight is relevant to our understanding of disease progression and cognitive symptomatology.

**Keypoints:**

- Regional activity and network clustering are pathologically high in glioma
- However, high-activity regions show low clustering and vice versa
- This finding could be relevant to understand functioning and prognosis in glioma

**Importance of the study:** Glioma patients show high peritumoral brain activity, which relates to faster tumor progression. Moreover, patients have local and global functional network disturbances, which associate with cognitive dysfunction and other symptoms. However, how such activity and network deviations correlate across and within patients is unclear. We, therefore, studied a large cohort of newly diagnosed glioma patients and matched healthy controls, extracting activity and connectivity from the entire cortex. We find a surprising relationship between deviations in activity and local clustering: while higher activity and clustering go hand in hand in controls, the pathologically high activity we observe in individual glioma patients coincides with exceedingly low clustering, while areas with normal activity levels have pathologically high clustering. These insights indicate an intricate relationship between aberrant activity and connectivity throughout the brain in glioma. It remains to be seen how this complex relationship impacts tumor growth and potentially cognitive deficits.

## Introduction

Glioma is the most common type of primary brain tumor. The prognosis is poor, and many patients suffer from debilitating symptoms such as cognitive dysfunction. In addition, patients show differences in whole-brain neurophysiology compared to healthy controls (HCs). Specifically, disturbances in neuronal activity and functional brain network connectivity have been found locally as well as throughout the brain. However, it is unclear how these indices of brain functioning relate to each other, while the interplay between activity and connectivity might be essential for prognosis and patient functioning.

In preclinical studies, glioma cells directly and reciprocally interact with their immediate neuronal environment.^1,2^ Via the formation of glutamatergic neuron-to-glioma synapses, neuronal spiking activity in the tumor’s proximity directly promotes tumor proliferation and invasion.^2,3^ Translational studies have used magnetoencephalography (MEG) as a non-invasive measurement of neuronal activity, reporting high activity around the tumor and across the tumor hemisphere as compared to controls.^4^ This pathologically high brain activity relates to shorter progression-free survival,^5,6^ even when using global brain activity measures, further underlining the clinical relevance of activity throughout the brain for tumor growth in glioma patients.

Glioma patients also show different activity synchronization between brain regions (i.e., functional connectivity) compared to healthy people. Functional connectivity is the statistical dependency between time series (activity patterns) from different brain regions.^7^ Based on the full matrix of pairwise connectivities between brain regions, network (graph) theory can be used to extract meaningful network topological markers.^8,9^ These measures can reflect whole-brain or regional topology. Typically, a combination of segregative and integrative topological measures are used to characterize the brain network, as the combination of local specialization and overall integration is considered essential for network functioning.^10^ Glioma patients show higher local, segregative connectivity, while integrative connectivity is lower in comparison to controls.^10–13^ As is the case for activity, higher functional connectivity is associated with shorter survival.^14^ Moreover, pathologically high global clustering, describing the overall segregative properties of the network,^8,9^ relates to poorer cognitive performance.^12,13,15,16^ Interestingly, these disturbances in local clustering are not limited to the peritumoral region: they do not correlate with distance from the tumor and are thought to represent a truly global network pathology.^12^

In support of the idea that the interaction between the ‘rest of the brain’ and glioma is complex and clinically important, we recently showed that gliomas tend to occur in regions with intrinsically higher brain activity in HCs.^17^ Moreover, while most tumors seem to occur in regions with intrinsically high clustering and integrative connectivity in controls,^12,18,19^ patients with gliomas located in regions with intrinsically low clustering have more extensively different functional network clustering at diagnosis.^12^ However, it is unclear how activity and connectivity are interrelated throughout the brain. Answering this question could help guide our thinking on tumor-brain cross-talk and its impact on disease progression and (management of) symptomatology.

Therefore, we focused on how these two aspects of neurophysiological functioning co-occur regionally, not only in the tumor region but throughout the brain, by collecting MEG in glioma patients and controls. We hypothesized that high local activity would relate to high local clustering, as local activity as measured with MEG already reflects the synchronous activity of large groups of neurons and may therefore imply local clustering.^20^ Furthermore, brain regions with higher levels of neuronal spiking activity also show higher integrative connectivity in (computational) studies.^21–23^ We thus hypothesized positive correlations between activity and connectivity, at least in HCs. Finally, we explored whether the level of peritumoral activity would drive the relationships between distant activity and network connectivity, which could indicate global effects of local pathological activity.

## Materials and Methods

### Participants

Patient data stemmed from an ongoing study at the Amsterdam UMC location VUmc that included newly diagnosed patients with diffuse glioma and has been published on in the past (Supplementary Table S1). Inclusion criteria were suspected glioma of grade II or higher as defined by the 2006 WHO classification,^24^ and age >18 years. Exclusion criteria were neuropsychiatric disorders or comorbidities of the central nervous system. We analyzed preoperative data only. For a posthoc test on the subtypes of glioma, we used molecular tumor markers that were established as part of routine clinical care after 2016 and the 2021 WHO classification was used to classify patients.^25^

A cohort of HCs was measured as part of two studies using the same MEG system and procedures,^26,27^ out of which we selected HCs matching patients’ sex and age at the group-level.

We investigated activity and network connectivity at three spatial levels in patients: (1) the area containing the tumor (peritumoral area), (2) its contralateral homologue, and (3) all areas that did not contain tumor (including contralateral homologue area). The main focus of this study was the regions that were not infiltrated by tumor on MRI, i.e. the rest of the brain (3),but results for the other two spatial levels (peritumoral area and contralateral homologue area) can be found in Supplementary Table S2. To define these peritumoral regions, tumor masks were either manually drawn in, slice by slice [LD], on post-gadolinium T1-weighted and FLAIR anatomical images,^28^ or automatically segmented using a neural network algorithm and visually checked.^29^ Using all 210 cortical regions of the Brainnetome atlas^30^, individual regions were considered part of the peritumoral area when at least 12% of the region’s volume overlapped with the tumor mask (Supplementary materials for more information). The contralateral homologue of the peritumoral area (2) was defined as the same atlas region(s) where the tumor was located but in the contralateral hemisphere. Patients with bilateral tumors or tumors that did not overlap for more than 12% with any region were excluded from analyses concerning the peritumoral and homologue areas. The rest of the brain included all regions that were not in the peritumoral area and did not include any tumor (0% overlap).

The VUmc Medical Ethics Committee approved all studies, and our research conduct followed the Declaration of Helsinki. Participants gave written informed consent before participation.

### Magnetoencephalography

Participants underwent a 5-minute eyes-closed resting-state MEG in supine position, using a 306-channel Elekta Neuromag Oy MEG (Helsinki, Finland) system in a magnetically shielded room, with a sampling frequency of 1250 Hz and online 0.1Hz high pass and 410Hz antialiasing filters. We used cross-validation signal space separation (SSS), after which raw data were visually inspected and malfunctioning channels were excluded [LD]. To remove artefacts offline, these channels were removed before applying the temporal extension of Signal Space Separation in MaxFilter software (Elekta NeuroMag Oy, version 2.2.15) to the raw data. The signal was subsequently filtered between 0.5-45Hz using a single-pass finite impulse response filter in MaxFilter. We used a 3D digitizer (Fastrak; Polhelmus, Colchester, VT, USA) to digitize 4 or 5 head position indicator coils, as well as the scalp surface and nose to enable co-registration the MEG data to the patients’ anatomical MRIs using surface matching. Subsequently, a scalar beamformer implementation (Elekta Neuromag Oy, version 2.1.28) source-reconstructed^31^ broadband (0.5-45Hz) MEG time series to the centroids^27^ of the 210 cortical regions of the Brainnetome atlas.^30^ We then selected 15 epochs for patients and eight epochs for HCs (each 4 * 4096 samples of 3.27s) for further analysis. These were the smallest number of good quality epochs available for any subject in the cohorts.

### Regional brain activity

The offset of the aperiodic part of the power spectrum was used as a proxy for neuronal spiking activity.^32,33^ Power spectra were obtained using Welch’s method with a hamming window for each epoch and cortical brain region. These spectra were averaged over all epochs per subject to obtain one spectrum per brain region. Finally, the Python implementation of the Fitting Oscillations & One Over F (FOOOF) toolbox (https://github.com/fooof-tools/fooof^32^) was used to estimate the offset by fitting the non-oscillatory part of the power spectrum using the exponential function L: L = b – log(k + F^x^). The parameter b is the offset, describing the power of the lowest frequency of the power spectrum; k is responsible for the bending of the aperiodic part and was set to 0. F is a vector containing all frequencies, and x is the slope of the aperiodic part.

In order to compare regional offset between patients and HCs, we standardizedd patients’ and HCs’ regional values based on the regional mean and standard deviation of HCs (Figure 1, Panel A visualizes the standardisation procedure). This allowed us to filter out intrinsic regional variations, leaving us with values representing deviations from HCs, hereafter referred to as ‘dev’ (e.g. offset_dev_) in this study.

**Figure 1.** **A** Visual representation of the standardisation procedure to obtain regional deviation values for offset, CC and EC. We filtered out premorbid regional variation in these measures using the regional mean and standard deviation of HCs. **B** Conceptual representation of the group-level analysis. For both patients and HCs we obtained the regional means of the raw offset, EC and CC values. We then performed the spin-test^37^ to relate offset to EC and CC on the group-level. **C** Conceptual representation of the within-subject analysis, for which we used LMMs to relate deviations in offset and network measures in the rest of the brain to each other.

### Functional networks

Functional networks were constructed in Python (version 3.19.5). We estimated the functional brain network by first employing a fast Fourier transform-based band-pass filter to every epoch per brain region. Functional networks were reconstructed for the delta (0.5-4Hz), theta (4-8Hz), and lower alpha (8-10Hz) bands since previous studies in glioma have predominantly found network alterations for these frequency bands.^10,15^ We used the Phase Lag Index (PLI) to calculate functional connectivity between all 210 cortical regions,^34^ as it has been amply used to establish functional networks in glioma.^12,13,15^ PLIs were calculated per epoch and subsequently averaged over epochs per frequency band. We then thresholded and binarized frequency-specific networks using a proportional threshold, keeping only the strongest 20% or 30% of connections, yielding six networks per participant (two densities for three frequency bands, Supplementary materials for additional information).

We calculated the local clustering coefficient (CC) and eigenvector centrality (EC) for all 210 regions using the Networkx Python package (version 2.3).^35^ The local CC is based on the connectivity between regions that a region is connected to, specifically the number of triangles formed between such regions.^9^ It is thought to represent segregative, ‘local’ (in terms of the network) connectivity. EC reflects the integrative properties of a node.^9^ It is based on the number of connections of the node itself and takes the number of connections of its neighbors and neighbor’s neighbors etc., into account.^36^

All CC and EC values were standardizedd based on regional means and standard deviations of HCs, to represent the deviation from controls (CC_dev_, EC_dev_).

### Statistical Analysis

All statistical analyses were performed in Python. To match the patients to the HCs, we used Mann-Whitney U and Chi-square tests to check for differences in age and sex between the two groups.

Differences in offset_dev_, local CC_dev_ and EC_dev_ between patients’ peritumoral, homologue and rest of the brain values and HCs whole-brain values_dev_ were calculated using the Mann-Whitney U test. To test whether peritumoral and contralateral homologue areas differed within patients, Wilcoxon signed-rank tests were used. All tests were performed for two network densities per three frequency bands.

Next, we explored the relationship between regional activity and functional network connectivity at the group-level (Figure 1, Panel B). We averaged *raw* offset, EC and CC values for every region over all participants per group, obtaining one value per region for the three measures. We then correlated offset with EC and CC (for two densities in three bands) using the spin test with a Pearson’s correlation implementation and 5000 permutations (https://www.github.com/spin-test).^37^ We used this test because regions located close together potentially show similar properties (e.g. activity), which might drive any spatial correlation between two variables. The spin test randomly rotates the spherical projections of all regional values and calculates the correlation an *n* number of times. The original correlation of interest can then be tested against these null models. We used binomial confidence intervals for the p-values to determine the significance of these correlations.

We then focused on within-subject effects (Figure 1, Panel C), using standardizedd values only. We used linear mixed models (LMMs) to handle within-subject dependencies between regions (Supplementary materials for an alternative approach using Pearson’s correlations). We fitted a model with offset_dev_ as the independent variable and CC_dev_ and EC_dev_ as predictors and included a random intercept for participants, using the statsmodels implementation (version 0.13.2) in Python. We fitted a separate model for every frequency band, density and group, yielding 12 models (3*2*2) in total. In another six models, group differences were tested through interaction terms (group x CC_dev_/EC_dev_). We reran this analysis using standardizedd values to obtain the standardizedd beta coefficients as an effect size metric. All of the analyses on the relationship between metrics focused on the rest of the brain of patients, excluding the (peri)tumoral region.

Lastly, using Pearson’s correlations, we tested whether peritumoral offset_dev_ was associated with the correlations (within-subject correlations, Supplementary materials) between activity and regional network characteristics throughout the brain. We used the mean offset_dev_ across all peritumoral regions and the mean of the three peritumoral regions with the highest offset_dev_ to be maximally sensitive to any effects.

All p-values were adjusted for multiple comparisons (across frequency bands and densities) using the false discovery rate (FDR^38^) and were deemed significant at adjusted p < 0.05 (*p*_*FDR*_). Results that were replicated for both network densities (20%, 30%) were deemed robust.

## Results

### Participant characteristics

MEGs of 84 glioma patients and 61 HCs were analyzed (Table 1 for participant characteristics).

**Table 1.** Participant characteristics

| Characteristics | Healthy controls (N = 61) | Glioma patients (N = 84) |
| --- | --- | --- |
| Age (mean (SD)) | 48.03 (9.62) | 45.68 (15.23) |
| Sex (number of females (males)) | 27 (34) | 23 (61) |
| Tumor WHO grade (II/III/IV) | NA | 37/30/17 |
| Tumor histology (GBM/A/O/NA) | NA | 30/30/23/1 |
| Tumor volume, corrected for headsize (mean ml (SD)) | NA | 39.8 (38.0) |
| Tumor side (left/right/bilateral) | NA | 49/31/4 |
| IDH-mutant, 1p/19q non-codeleted glioma (number (%)) | NA | 28 (33.3) |
| IDH-mutant, 1p/19q-codeleted glioma (number (%)) | NA | 17 (20.2) |
| IDH-wildtype glioblastoma (number (%)) | NA | 30 (35.7) |
| Unknown molecular subtype (number (%)) | NA | 9 (10.7) |
| Epilepsy (yes (no)) | NA | 70 (14) |
| KPS (median (range) / NA) | NA | 100 (50 - 100) / 11 |

*Note.* SD = Standard Deviation, GBM = Glioblastoma, A = Astrocytoma, O = Oligodendroglioma, NA = Not available

### Higher activity and clustering in patients

Patients’ offset_dev_ was significantly higher than controls’ values in all areas: the peritumoral area (mean = 1.559, SD = 1.513, *U* = 3554, *p*_*FDR*_ *= <*.*001*), contralateral homologue area (mean = 0.373, SD = 1.168, *U* = 2616, *p*_*FDR*_ = .006) and rest of the brain (mean = 0.375, SD = 1.269, *U* = 3319, *p*_*FDR*_ = .004), suggesting that brain activity is pathologically high throughout the brain in glioma patients (Figure 2A). Furthermore, within patients, peritumoral offset_dev_ was higher than its contralateral homologue (*Z* = 133, p < .001).

**Figure 2.** **A** Offset in peritumoral, homologue and non-tumoral (rest of the brain) areas in glioma patients and the whole brain of HCs. **B** Delta, theta and lower alpha band local CC (higher panel) and EC (lower panel) in patients (rest of the brain) and HCs (at 30% density of the network). **C** Within-subject relations between offset_dev_ and lower alpha band CC_dev_ (higher panel) and EC_dev_ (lower panel). The left column shows the correlations plotted for the different subgroups investigated. The right column shows the relationship between offset_dev_ and and CC_dev_ (higher panel) and EC_dev_ (lower panel) with a regression line and confidence interval (shaded region) drawn based on all data points. The y-axis represents offset_dev_ while the x-axis represents the network metrics. All relationships are plotted for the rest of the brain of patients for the lower alpha band and 30% density of the network. For all three subplots, we used raincloud plots, consisting of a half-density plot, a boxplot and a scatter plot, to visualize the findings. This type of visualization enables us to show several aspects of the data that would be lost if only using one of these three modalities. The half density plot visualizes the distribution of datapoints, while the boxplot enables us to show descriptive statistics at the same time. Finally the scatter shows the raw data points, giving us another insight in the distribution of the data. These plots were created with the help of the python implementation for raincloud plots (https://github.com/pog87/PtitPrince)^50^.

Compared to HCs, CC_dev_ was higher peritumorally (delta band), in the contralateral homologue (delta, lower alpha band) and the entire rest of the brain (all bands, Table 2, Table 3, Figure 2B). Delta band EC_dev_ was lower in patients in the rest of the brain but not in the peritumoral area or its homologue (Table 2, Table 3, Figure 2B). These results suggest that clustering is globally higher in glioma patients, while integrative connectivity is lower in the non-tumoral areas. CC_dev_ and EC_dev_ did not differ between the peritumoral area and its homologue within patients (Supplementary Table S3, Figure 2B).

**Table 2.**
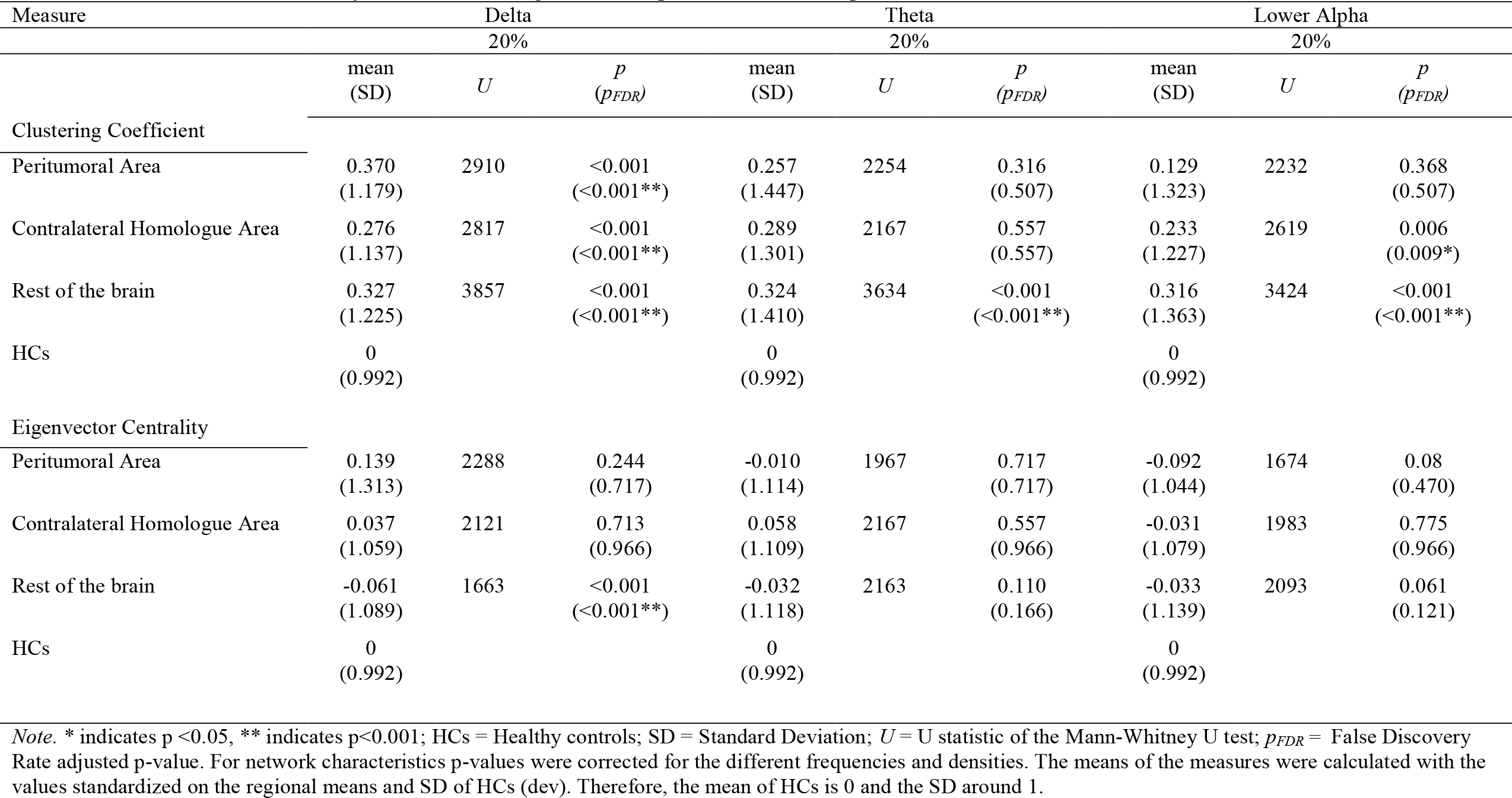
Network characteristics (density 20%) in the investigated areas of patients and their comparison to whole brain characteristics of HCs

**Table 3.** Network characteristics (density 30%) in the investigated areas of patients and their comparison to whole brain characteristics of HCs

| Measure | Delta |  |  | Theta |  |  | Lower Alpha |  |  |
| --- | --- | --- | --- | --- | --- | --- | --- | --- | --- |
|  | 30% |  |  | 30% |  |  | 30% |  |  |
|  | mean<br>(SD) | *U* | *p*<br>(*p<sub>FDR</sub>*) | mean<br>(SD) | *U* | *p*<br>(*p<sub>FDR</sub>*) | mean<br>(SD) | *U* | *p*<br>(*p<sub>FDR</sub>*) |
| **Clustering Coefficient** |  |  |  |  |  |  |  |  |  |
| Peritumoral Area | 0.364<br>(1.156) | 2920 | <0.001<br>(<0.001**) | 0.246<br>(1.466) | 2169 | 0.551<br>(0.551) | 0.155<br>(1.350) | 2212 | 0.423<br>(0.507) |
| Contralateral Homologue Area | 0.335<br>(1.159) | 2906 | <0.001<br>(<0.001**) | 0.297<br>(1.320) | 2295 | 0.231<br>(0.277) | 0.225<br>(1.209) | 2670 | 0.003<br>(0.005*) |
| Rest of the brain | 0.399<br>(1.231) | 3948 | <0.001<br>(<0.001**) | 0.351<br>(1.377) | 3711 | <0.001<br>(<0.001**) | 0.295<br>(1.290) | 3390 | <0.001<br>(<0.001**) |
| HCs | 0<br>(0.992) |  |  | 0<br>(0.992) |  |  | 0<br>(0.992) |  |  |
| **Eigenvector Centrality** |  |  |  |  |  |  |  |  |  |
| Peritumoral Area | 0.128<br>(1.305) | 2206 | 0.439<br>(0.717) | -0.017<br>(1.098) | 2163 | 0.570<br>(0.717) | -0.036<br>(1.030) | 1944 | 0.637<br>(0.717) |
| Contralateral Homologue Area | 0.035<br>(1.079) | 2020 | 0.913<br>(0.966) | 0.072<br>(1.097) | 1972 | 0.735<br>(0.966) | -0.025<br>(1.046) | 2034 | 0.966<br>(0.966) |
| Rest of the brain | -0.058<br>(1.106) | 1582 | <0.001<br>(<0.001**) | -0.023<br>(1.122) | 2208 | 0.157<br>(0.188) | -0.019<br>(1.127) | 2308 | 0.310<br>(0.310) |
| HCs | 0<br>(0.992) |  |  | 0<br>(0.992) |  |  | 0<br>(0.992) |  |  |

*Note.* \* indicates $p < 0.05$ , \*\* indicates $p < 0.001$ ; HCs = Healthy controls; SD = Standard Deviation; *U* = U statistic of the Mann-Whitney U test; $p_{FDR}$ = False Discovery Rate adjusted p-value. For network characteristics p-values were corrected for the different frequencies and densities. The means of the measures were calculated with the values standardized on the regional means and SD of HCs (dev). Therefore, the mean of HCs is 0 and the SD around 1.

A posthoc test revealed similar profiles for the different subtypes of glioma (Supplementary materials and Supplementary Table S4, Table S5, Table S6).

### Positive group-level regional correlations

Raw offset values related positively to clustering across frequencies in HCs and patients, while it related positively to theta band EC in HCs, but not patients (Supplementary Table S7).

### Negative within-subject regional correlations

With respect to clustering, LMMs revealed that within patients, regional offset_dev_ related negatively to regional lower alpha band CC_dev_ in the rest of the brain of patients (Table 4, Figure 2C). This relationship differed significantly from that in HCs (Supplementary Table S8), where no significant associations were found for the lower alpha band (Supplementary Table S9). HCs did show a positive relationship between offset and CC in the delta band (Supplementary Table S9).

**Table 4.** Linear mixed model with offset_dev_ as dependent and EC_dev_ and CC_dev_ as independent variables for the rest of the brain of patients

| Frequency,<br>Density | Variable | Coefficient [CI] | Std coefficient<br>(beta) | Z | p | $p_{FDR}$ |
| --- | --- | --- | --- | --- | --- | --- |
| **Delta** |  |  |  |  |  |  |
| 20% | Intercept | 0.383 [0.225, 0.542] |  | 4.734 | <0.001 |  |
| | $\text{EC}_{\text{dev}}$ | 0.005 [-0.010, 0.020] | 0.004 | 0.681 | 0.496 | 0.541 |
| | $\text{CC}_{\text{dev}}$ | 0.011 [-0.003, 0.025] | 0.011 | 1.541 | 0.123 | 0.211 |
| 30% | Intercept | 0.381 [0.223, 0.540] |  | 4.708 | <0.001 |  |
| | $\text{EC}_{\text{dev}}$ | -0.001 [-0.016, 0.013] | -0.001 | -0.189 | 0.850 | 0.850 |
| | $\text{CC}_{\text{dev}}$ | 0.013 [-0.001, 0.027] | 0.013 | 1.85 | 0.064 | 0.150 |
| **Theta** |  |  |  |  |  |  |
| 20% | Intercept | 0.385 [0.226, 0.544] |  | 4.75 | <0.001 |  |
| | $\text{EC}_{\text{dev}}$ | 0.013 [-0.001, 0.028] | 0.012 | 1.778 | 0.075 | 0.150 |
| | $\text{CC}_{\text{dev}}$ | 0.007 [-0.007, 0.020] | 0.007 | 0.984 | 0.325 | 0.390 |
| 30% | Intercept | 0.384 [0.225, 0.543] |  | 4.739 | <0.001 |  |
| | $\text{EC}_{\text{dev}}$ | 0.011 [-0.004, 0.025] | 0.009 | 1.436 | 0.151 | 0.227 |
| | $\text{CC}_{\text{dev}}$ | 0.008 [-0.006, 0.022] | 0.008 | 1.111 | 0.267 | 0.356 |
| **Lower Alpha** |  |  |  |  |  |  |
| 20% | Intercept | 0.392 [0.234, 0.551] |  | 4.845 | <0.001 |  |
| | $\text{EC}_{\text{dev}}$ | -0.063 [-0.077, -0.048] | -0.056 | -8.349 | <0.001 | <0.001** |
| | $\text{CC}_{\text{dev}}$ | -0.025 [-0.039, -0.012] | -0.027 | -3.636 | <0.001 | <0.001** |
| 30% | Intercept | 0.398 [0.240, 0.557] |  | 4.922 | <0.001 |  |
| | $\text{EC}_{\text{dev}}$ | -0.056 [-0.071, -0.042] | -0.05 | -7.564 | <0.001 | <0.001** |
| | $\text{CC}_{\text{dev}}$ | -0.044 [-0.058, -0.030] | -0.044 | -6.103 | <0.001 | <0.001** |

Note. \* indicates $p < 0.05$ , \*\* indicates $p < 0.001$ ; A random intercept was fitted for participants; CI = Confidence interval for coefficient; Std = Standardized; $p_{FDR}$ = False Discovery Rate adjusted p-value. The p-values were corrected for the different frequency bands and densities. Only the independent variables were included in this correction.

These results counterintuitively indicate that in patients, regionally, pathologically high offset associates with lower deviating CC, even though our previous results established pathologically high offset and CC in patients throughout the brain. As can be seen in Figure 2C, offset_dev_ values that were more similar to HCs (around 0) were associated with pathologically high CC_dev_.

Similarly, offset_dev_ related negatively to lower alpha band EC_dev_ (Table 4, Figure 2C). Again, there was a significant difference between patients and HCs in this relationship (Supplementary Table S8), with HCs not showing an association between offset_dev_ and Ec_dev_ in the lower alpha band (Supplementary Table S9). Conversely, HCs showed a positive relationship between offset_dev_ and EC_dev_ in the delta band (Supplementary Table S9), which differed significantly from that in patients (Supplementary Table S8).

As a second approach to investigate the within-subject relationships, we used Pearson correlations, which yielded similar results as the LMMs (Supplementary materials Table S10). In addition, a posthoc test revealed that these negative relationships were predominantly present in IDH-wildtype glioblastomas (for the relationship between CC_dev_, EC_dev_ and offset_dev_) and IDH-mutant, 1p/19q codeleted gliomas (for the relationship between EC_dev_ and offset_dev_, Supplementary materials Table S11, Table S12).

### Activity-dependence of regional correlations

We found no significant associations between peritumoral offset_dev_ and the within-patient associations between regional offset_dev_ and either CC_dev_ or EC_dev_ (Supplementary Table S13), suggesting that the negative correlations between activity and connectivity throughout the brain were not dependent on the level of peritumoral activity.

## Discussion

We investigated how regional brain activity and functional network connectivity relate to each other in patients with glioma and healthy subjects. We surprisingly found that although both brain activity and network clustering were significantly higher throughout the brains of glioma patients compared to controls, non-tumoral regional deviations in activity and clustering correlate negatively within patients. In other words, regions marked by pathologically high brain activity typically show very low network clustering compared to controls, while areas with regular brain activity have very high network clustering. The regions with normal clustering are between these extremes, which display only slightly higher activity than controls.

As expected, brain activity was higher around the tumor, which aligns with others’ and our previous work.^2,4^ We may speculate that around the tumor, heightened activity is driven by reciprocal neuron-glioma cell interactions,^1,2^ by increased glutamate being present around the tumor,^40^ or both. The mechanisms leading to heightened activity further away from the tumor, as we find here and in our previous work,^2,4^ remain elusive. Do invasive glioma cells form neuron-glioma synapses far away from the tumor? Does high activity in the peritumoral region somehow ‘spread’ to other brain areas?

We also observed deviant functional network connectivity in patients, as expected. Local clustering, representing segregative connectivity throughout the brain, was higher in patients than in controls. Several studies, some using data from the same cohort, align with this finding, particularly for the theta band.^12,13,15,16^ Furthermore, patients’ regional centrality, a measure of integrative connectivity, was lower compared to controls for the delta band, in line with previous work.^10^ Overall, these observations corroborate previous MEG and functional MRI studies, suggesting that local, segregative connectivity is higher while integrative connectivity is lower in glioma.^10,11,13,41–43^

But what mechanisms could be at the core of such widespread network disturbances in segregative and integrative connectivity? We could speculate that the growing tumor initially has a local impact on the functional network, which subsequently affects the network on a larger scale, seeing as other regions ‘take over’ some functioning from locally affected regions, as postulated in the cascadic network failure model.^44^ On a more cellular level, *invasive* glioma cells might directly affect the functional network topology throughout the brain. Glioma, and especially the aggressive IDH-wildtype glioma (glioblastoma), are marked by highly invasive cells that infiltrate the surrounding brain via white matter tracts, blood vessels, and microtubules, beyond the area where the mass of the tumor is located. There, these invasive cells adhere to other cells, such as neurons.^45^ Potentially, the invasive cells may spread in the brain and impact the local environment and functioning of neuronal cell populations further away from the mass of the tumor, thereby impacting functional network dynamics throughout the brain. Especially clustering is typically a local network process (although the measure itself is not based on anatomical (Euclidean) distance and could thus also pick up connectivity between functionally connected but spatially distant regions), and such local cellular dynamics might regionally disturb the clustering in regions further away from the tumor. However, the extent of neurogliomal synapses commonly formed at locations distant from the tumor is unknown. As such, the exact mechanisms underlying the observed functional network differences remain to be elucidated; we might expect that several mechanisms, for instance cellular invasion and cascadic network failure, are at play simultaneously. Longitudinal studies investigating these processes on multiple scales are warranted.

Our findings confirm the hypothesis that higher regional brain activity generally relates to higher connectivity, as seen in our group-level results performed on raw values instead of deviations from the controls. Indeed, few studies have investigated the relationship between brain activity and connectivity and found that they positively relate to each other in the healthy setting.^21–23^

Interestingly, our within-subject analyses revealed a different relationship between activity and patient connectivity deviations. When zooming in on the within-subject level, an interesting pattern emerged: deviations in brain activity and lower alpha clustering related negatively, indicating that pathologically high brain activity went hand in hand with very low clustering across regions within the same patient. This was particularly surprising as we found both brain activity and clustering to be higher throughout the brains of patients. This negative relationship may indicate that different regions showed these two types of neurophysiological deviations. Based on our findings, we may speculate that regions with the highest activity, as we show in the group comparisons, have pathologically low levels of clustering. Conversely, regions with more typical activity levels are either normal or very high regarding their level of clustering. This study was cross-sectional, so we cannot draw clear conclusions on the chronological emergence of deviations in clustering and brain activity. However, we may hypothesize that in regions where brain activity is highest, intrinsic oscillatory patterns might be altered such that it disconnects from regions that it is normally connected to. For regions showing activity that is similar to HCs but very high clustering, we may postulate that this pattern of deviations has a protective nature: maybe high clustering helps to maintain more normal levels of activity throughout the brain by ‘breaking up’ the functional network. Such a scenario has been posited in epilepsy previously, as epilepsy patients show less integration and heightened segregation of the functional network in the interictal period.^46^ The epileptic zone seems to be functionally isolated from other regions through higher connectivity within itself and lower centrality.^47,48^ It has been speculated that such isolation and breaking up of the functional network might lower susceptibility to new seizures.^47,49^ This mechanism may also play a role in our glioma patients, particularly since almost all suffer epileptic seizures.

Regarding centrality, we similarly found a negative relationship between brain activity and centrality in the lower alpha band. This finding was less surprising, as we did observe centrality to be lower throughout the brain in the patient group, but only for the delta band. This negative relationship might indicate that regions showing the most pathologically high activity exhibit the lowest levels of centrality (in the lower alpha band) and vice versa. This is relevant in the context of the cascadic network failure model, where regions that are very central to the network take over functions from other regions that are affected by the lesion.^44^ Such taking over of activity is thought to cause the central region to overload and eventually fail as a hub in the network. Speculatively, we may interpret that such failing of regions is reflected in the current results: regions with the highest level of activity failed to be central integrators and showed the lowest level of centrality, even though we cannot establish the timeline of activity and centrality changes in this study.

Another issue to consider is that peritumoral activity is pathologically high in glioma, while glioma is also known to occur most often in regions that are intrinsically high in activity and connectivity in controls.^18,19^ Based on our cross-sectional data, we cannot disentangle whether high-activity regions in glioma patients were premorbidly already highly active or became pathological upon glioma occurrence. Furthermore, we found lower EC in patients only for the delta band but found a negative relationship with activity for the lower alpha bands. As our results are frequency dependent, this warrants caution in the interpretations of these results.

Finally, we explored whether higher peritumoral brain activity affects the relationships between deviations in activity and network characteristics. Our analysis showed that the level of brain activity in the peritumoral area did not relate to the observed relationships. This suggests that local, aberrant activity around the tumor is likely not the primary driver of network-activity correlations further away in this cohort but that more complex mechanisms are at play.

This study has several limitations. Firstly, the study was cross-sectional and correlational, making it difficult to derive firm conclusions about the mechanisms underlying the observed disturbances and their interplay. Studying this longitudinally before tumor resection would be ideal for understanding how deviations develop over time. However, these data are almost impossible to collect in this patient population, as treatment starts as soon as possible. A second limitation of the current study is the small correlation values and coefficients that we observed, particularly in the within-subject analyses. We tried to counter this by employing different types of analyses (Supplementary materials) and ways to threshold the functional networks to test the robustness of results, allowing us to draw cautious conclusions. Also, specific preprocessing choices may affect the activity measures used in this study. For example, the lowest frequency that can be measured from the MEG signal determines the fitting of the FOOOF model and the offset that is captured. Therefore, specific choices in the preprocessing and specific filtering of the MEG signal before analysis affect the offset that can be measured. Future studies should focus on further exploring the parameter space and how specific preprocessing choices may affect results. The same holds for choices in the construction of the connectivity matrix and thresholding and binarisation of the constructed functional networks.

## Conclusion

While brain activity and local clustering are pathologically high throughout the brain in glioma patients, regionally, these neurophysiological deviations present in a complex manner. Regions with the highest activity show lower-than-normal clustering. Future studies should focus on further characterising these whole-brain deviations and their development over time. This might aid in understanding such disturbances better and uncover how neuron-glioma interactions shape clinical functioning and influence prognosis.

## Data Availability

All scripts used to analyse the data can be openly accessed on our github repository: https://github.com/multinetlab-amsterdam/projects/tree/master/activity_network_project_2023

## Conflict of Interest

Mona LM Zimmermann: None declared

Lucas C Breedt: None declared

Eduarda GZ Centeno: None declared

Jaap C Reijneveld:

Payment for educational event: iMeedu Teaching course ‘Noord-Noordhollandse

Neurologen’, iMeedu Teaching course ‘Het Centrale Brein’

Support for attending meetings or travel: Arvelle Therapeutics (manufacturer of anti-seizure medication Cenobamate), attended European Epilepsy Congress in Geneva, 2023

Fernando AN Santos: None declared

Cornelis J Stam: None declared

Marike R van Lingen: Grants or contracts from any entity: MEGIN

Menno M Schoonheim:

Grants or contract from any entity: Dutch MS foundation, Eurostars-EUREKA, ARSEP, Amsterdam Neuroscience, MAGNIMS, ZonMW (Vidi grant 09150172010056)

Consulting fees: Atara Biotherapeutics, Biogen, Celgene/Bristol Meyers Squibb, Genzyme, MedDay, Merck

On the editorial board of: Neurology and Frontiers in Neurology

Arjan Hillebrand:

Grants or contracts from any entity: MEGIN, KWF Dutch Cancer Society

Linda Douw:

Grants or contracts from any entity: MEGIN

## Authorship

Design and conceptualization of study: M.L.M.Z, L.D, J.C.R, F.A.N.S, L.C.B, F.A.N.S, M.R.vL

Data collection and preprocessing: M.R.vL, L.C.B, A.H., M.M.S, C.J.S

Data analysis: M.L.M.Z, L.D, E.G.Z.C, L.C.B, F.A.N.S

Interpretation of results: All authors

Writing of the manuscript: M.L.M.Z

Revision and final approval of the manuscript: All authors

## Data Availability

Data will be made available under reasonable request. All scripts used to analyse the data can be openly accessed on our github repository: https://github.com/multinetlab-amsterdam/projects/tree/master/activity_network_project_2023

