## Supplementary materials for "The relationship between pathological brain activity and functional network connectivity in glioma patients"

**Table S1 Overview of studies that published on (partly) the same data as the current study**

| Authors (Year) | Title |
|---|---|
| Douw et al. (2010)[1] | Epilepsy is related to theta band brain connectivity and network topology in brain tumor patients |
| van Dellen et al. (2012)[2] | MEG Network Differences between Low- and High-Grade Glioma Related to Epilepsy and Cognition |
| van Dellen et al. (2012)[3] | Connectivity in MEG resting-state networks increases after resective surgery for low-grade glioma and correlates with improved cognitive performance. |
| Carbo et al. (2017)[4] | Dynamic hub load predicts cognitive decline after resective neurosurgery |
| Derks et al. (2018)[5] | Oscillatory brain activity associates with neuroligin-3 expression and predicts progression free survival in patients with diffuse glioma. |
| Derks et al. (2019)[6] | Understanding cognitive functioning in glioma patients: The relevance of IDH-mutation status and functional connectivity |
| Belgers et al. (2020)[7] | Postoperative oscillatory brain activity as an add-on prognostic marker in diffuse glioma. |
| Numan et al. (2021)[8] | Non-invasively measured brain activity and radiological progression in diffuse glioma |
| Derks et al. (2021)[9] | Understanding Global Brain Network Alterations in Glioma Patients |
| Röttgering et al. (2022)[10] | Toward unravelling the correlates of fatigue in glioma |
| Röttgering et al. (2023)[11] | Symptom networks in glioma patients: understanding the multidimensionality of symptoms and quality of life |
| van Lingen et al. (2023) [12] | The longitudinal relation between executive functioning and multilayer network topology in glioma patients |
| *Note.* Table adapted from Röttgering et al. (2023)[11] | |

***Table S2*** Linear Mixed Model with $offset_{dev}$ as dependent and $EC_{dev}$ and $CC_{dev}$ as independent variables for the peritumoral and contralateral homologue areas in patients

| Frequency, Density | Variable | Coefficient [CI] | *Z* | *p* | $p_{FDR}$ |
|---|---|---|---|---|---|
| Peritumoral Area | | | | | |
| Delta | | | | | |
| 20% | Intercept | 1.573 [1.276, 1.871] | 10.358 | < 0.005 | |
| | $EC_{dev}$ | 0.073 [-0.005, 0.151] | 1.832 | 0.067 | 0.200 |
| | $CC_{dev}$ | 0.022 [-0.057, 0.102] | 0.549 | 0.583 | 0.777 |
| 30% | Intercept | 1.546 [1.249, 1.843] | 10.192 | <0.001 | |
| | $EC_{dev}$ | 0.078 [-0.001, 0.156] | 1.945 | 0.052 | 0.200 |
| | $CC_{dev}$ | 0.083 [0.003, 0.163] | 2.022 | 0.043 | 0.200 |
| Theta | | | | | |
| 20% | Intercept | 1.591 [1.291, 1.891] | 10.397 | <0.005 | |
| | $EC_{dev}$ | 0.089 [0.007, 0.171] | 2.121 | 0.034 | 0.200 |
| | $CC_{dev}$ | 0.013 [-0.062, 0.087] | 0.329 | 0.742 | 0.815 |
| 30% | Intercept | 1.586 [1.286, 1.886] | 10.372 | <0.005 | |
| | $EC_{dev}$ | 0.074 [-0.01, 0.157] | 1.728 | 0.084 | 0.201 |
| | $CC_{dev}$ | 0.035 [-0.041, 0.11] | 0.899 | 0.369 | 0.632 |
| Lower Alpha | | | | | |
| 20% | Intercept | 1.600 [1.301, 1.9] | 10.486 | <0.005 | |
| | $EC_{dev}$ | -0.011 [-0.104, 0.082] | -0.234 | 0.815 | 0.815 |
| | $CC_{dev}$ | -0.029 [-0.112, 0.053] | -0.698 | 0.485 | 0.727 |
| 30% | Intercept | 1.588 [1.289, 1.887] | 10.415 | <0.005 | |
| | $EC_{dev}$ | -0.049 [-0.139, 0.042] | -1.054 | 0.292 | 0.584 |
| | $CC_{dev}$ | 0.013 [ -0.069, 0.095] | 0.309 | 0.757 | 0.815 |
| Contralateral Homologue Area | | | | | |
| Delta | | | | | |
| 20% | Intercept | 0.398 [0.155, 0.642] | 3.203 | 0.001 | |
| | $EC_{dev}$ | -0.021[-0.087,0.044] | -0.633 | 0.527 | 0.584 |
| | $CC_{dev}$ | -0.038 [-0.097, 0.022] | -1.241 | 0.214 | 0.409 |
| 30% | Intercept | 0.392 [0.148, 0.635] | 3.155 | 0.002 | |
| | $EC_{dev}$ | -0.03 [-0.095, 0.035] | -0.902 | 0.367 | 0.489 |
| | $CC_{dev}$ | -0.019 [-0.077, 0.04] | -0.62 | 0.535 | 0.584 |
| Theta | | | | | |
| 20% | Intercept | 0.373 [0.135, 0.612] | 3.064 | 0.002 | |
| | $EC_{dev}$ | 0.08 [0.018, 0.612] | 2.532 | 0.011 | 0.045* |
| | $CC_{dev}$ | 0.032 [-0.025, 0.09] | 1.097 | 0.273 | 0.409 |
| 30% | Intercept | 0.379 [0.14, 0.618] | 3.108 | 0.002 | |
| | $EC_{dev}$ | 0.079 [0.018, 0.141] | 2.537 | 0.011 | 0.045* |
| | $CC_{dev}$ | 0.009 [-0.05, 0.068] | 0.303 | 0.762 | 0.540 |
| Lower Alpha | | | | | |
| 20% | Intercept | 0.364 [0.119, 0.609] | 2.915 | 0.004 | |
| | $EC_{dev}$ | 0.041 [-0.027, 0.109] | 1.175 | 0.240 | 0.409 |
| | $CC_{dev}$ | 0.053 [-0.008, 0.113] | 1.711 | 0.087 | 0.260 |
| 30% | Intercept | 0.353 [0.108, 0.598] | 2.827 | 0.005 | |
| | $EC_{dev}$ | 0.043 [-0.025, 0.111] | 1.242 | 0.214 | 0.409 |
| | $CC_{dev}$ | 0.084 [0.024, 0.145] | 2.736 | 0.006 | 0.045* |

*Note.* * indicates p <0.05, ** indicates p<0.001; A random intercept was fitted for participants; CI = Confidence interval for coefficient; $p_{FDR}$ = False Discovery Rate adjusted p-value. The p-values were corrected for the different frequency bands and densities, separately for the two areas. Only the independent variables were included in this correction.

### Tumor masks and area definitions

To define the peritumoral area, masks were either manually drawn in, slice by slice [LD], on post-gadolinium T1-weighted and FLAIR anatomical images,[13] or automatically segmented using a neural network algorithm.[14] Subsequently, for every subject, we calculated the volume overlap of the tumor masks with the regions of the BNA using FSL (version 6.0.5.1) to determine which BNA regions contained tumor. We then calculated the percentage overlap between the tumor and every region by dividing the normal volume of a region with the volume of the tumor mask within that region. Next, we plotted all percentage volume overlaps of all regions of all subjects in a histogram. This helped us to determine the percentage overlap that was the minimum overlap still commonly represented in patients. This minimum overlap was 12%. Therefore, regions were defined to be part of the peritumoral area when at least 12% of the region's volume overlapped with the tumor mask.

### Functional network thresholding

There is no agreed standard pipeline to threshold functional networks as of yet. We decided to use a proportional threshold by keeping only the n% strongest links. We used multiple densities (20%, 30%) to investigate whether results would replicate across densities and therefore be robust. At first, we additionally calculated a threshold of 10%. However, networks only containing the 10% strongest links showed many unconnected, isolated nodes, not allowing us to investigate our graph theoretical measures of interest. Therefore, we decided not to go further with a 10% threshold in our final analysis. We used the same thresholding procedure for all subjects (patients and HCs) in the study.

### Within-subject relationships using Pearson correlations

As a second approach to the within-subject analysis, we correlated regional $offset_{dev}$ with $CC_{dev}$ and $EC_{dev}$ using Pearson's correlation in every participant. To obtain two group-level within-subject correlation values, we calculated the weighted mean of the correlations by first z-transforming the correlations using Fisher's z- transform and then weighting these by the number of regions that were used in the initial correlation and finally taking the mean. To see whether this correlation was significant at the group-level, we used a Wilcoxon signed rank test against 0, in which we inputted each participant's Fisher z-transformed correlation value. To then test whether the relationship in patients differed from that in HCs, we used a Mann-Whitney U test with Fisher z-transformed correlations as input. Results from these analysis were similar to the LMM approach: $offset_{dev}$ related negatively to lower alpha $CC_{dev}$ in the rest of the brain of patients, but only for a 20% density after FDR correction (Table S10). This significantly differed from HCs for 30% density, who again did not show a relationship between $offset_{dev}$ and $CC_{dev}$ for the lower alpha band (Table S10). HCs again showed a positive relationship between delta $offset_{dev}$ and $CC_{dev}$, but this did not differ significantly from patients.

The relationship between $offset_{dev}$ and $EC_{dev}$ for the lower alpha band was similar to the LMMs when using Pearson's correlations for patients with $offset_{dev}$ relating negatively to $EC_{dev}$ (Table S10). For HCs, the positive relationship between $offset_{dev}$ and delta $EC_{dev}$, was similar as well and was now significantly different from that in patients (Table S10).

***Table S3*** Network characteristics in the investigated areas of patients and the comparison between peritumoral and contralateral homologue areas

| Measure, Area, Comparison | Delta | | Theta | | Lower Alpha | |
|---|---|---|---|---|---|---|
| | 20% | 30% | 20% | 30% | 20% | 30% |
| **Clustering Coefficient** | | | | | | |
| Peritumoral Area (mean (SD)) | 0.370 (1.179) | 0.364 (1.156) | 0.257 (1.447) | 0.246 (1.466) | 0.129 (1.323) | 0.155 (1.350) |
| Contralateral Homologue Area (mean (SD)) | 0.276 (1.137) | 0.335 (1.159) | 0.289 (1.301) | 0.297 (1.320) | 0.233 (1.227) | 0.225 (1.209) |
| Comparison Peritumoral and Contralateral Homologue Area ($U$, ($p$, $p_{FDR}$)) | 975 (0.306, 0.611) | 1134 (0.975, 0.975) | 1123 (0.920, 0.975) | 1132 (0.965, 0.975) | 862 (0.084, 0.501) | 955 (0.250, 0.611) |
| **Eigenvector Centrality** | | | | | | |
| Peritumoral Area (mean (SD)) | 0.139 (1.313) | 0.128 (1.305) | -0.010 (1.114) | -0.017 (1.098) | -0.092 (1.044) | -0.036 (1.030) |
| Contralateral Homologue Area (mean (SD)) | 0.037 (1.059) | 0.035 (1.079) | 0.058 (1.109) | 0.072 (1.097) | -0.031 (1.079) | -0.025 (1.046) |
| Comparison Peritumoral and Contralateral Homologue Area ($U$ ($p$, $p_{FDR}$)) | 1075 (0.689, 0.940) | 1080 (0.712, 0.940) | 1100 (0.808, 0.940) | 1127 (0.940, 0.940) | 978 (0.315, 0.940) | 1043 (0.549, 0.940) |

*Note.* * indicates p <0.05, ** indicates p<0.001; SD = Standard Deviation; $U$ = U statistic of the Mann-Whitney U test; $p_{FDR}$ = False Discovery Rate adjusted p-value. P-values were corrected for the different frequency bands and densities. The means of the measures were calculated with the values standardized on the regional means and SD of HCs (dev).

**Post-hoc subgroup analyses**

In order to better understand the surprising relationship between $CC_{dev}$ and $offset_{dev}$, we performed post-hoc analyses within patient subgroups according to molecular tumor types, namely IDH-wildtype glioblastoma, IDH-mutant, 1p/19q-codeleted, and IDH-mutant, 1p/19q non-codeleted glioma patients. All subgroups showed higher peritumoral activity in comparison to HCs (Table S4), while only patients with IDH-mutant, 1p/19q-codeleted and non-codeleted gliomas showed higher activity throughout the brain. Network characteristics per subgroup were similar to results from the entire group (Table S5, Table S6). The post-hoc tests of within-subject analysis focused on the lower alpha band, based on the interesting relationships between $CC_{dev}$ and $offset_{dev}$ that we found in the group-level analyses of this study. The negative correlation between $offset_{dev}$ and $CC_{dev}$ was significant in patients with an IDH-wildtype glioblastoma (only for one density) for LMMs (Table S11), but not when performing the correlation analysis (Table S12). The relationship between $offset_{dev}$ and $EC_{dev}$ was negative for IDH-wildtype glioblastoma and IDH-mutant, 1p/19q-codeleted gliomas, in both the LMM and correlation analyses (Table S11, S12). Finally, we again did not find a significant relationship between peritumoral offset and these correlations for either of the molecular subtypes, further indicating that the observed effects are widespread and independent of activity differences directly around the tumor (Table S13).

***Table S4*** Offset in the glioma subtypes for the investigated areas including comparison to HCs (whole brain) and between peritumoral and homologue areas

| Subtype | Peritumoral Area | | | Contralateral Homologue Area | | | Rest of the brain | | | Comparison Peritumoral Area to Homologue Area | |
|---|---|---|---|---|---|---|---|---|---|---|---|
| | mean (SD) | *U* | *p* (*p*$_{FDR}$) | mean (SD) | *U* | *p* (*p*$_{FDR}$) | mean (SD) | *U* | *p* (*p*$_{FDR}$) | *Z* | *p* |
| IDH-wildtype glioblastoma | 1.841 (1.579) | 1309 | <0.0001 (<0.001**) | 0.292 (1.132) | 864 | 0.199 (0.199) | 0.339 (1.349) | 1129 | 0.071 (0.071) | 7 | <0.001** |
| IDH- mutant, 1p/19q non-codeleted | 1.666 (1.622) | 1189 | <0.0001 (<0.001**) | 0.266 (1.166) | 863 | 0.106 (0.106) | 0.403 (1.174) | 1125 | 0.002 (0.048*) | 5 | <0.001** |
| IDH-mutant, 1p/19q-codeleted | 1.217 (1.302) | 565 | <0.001 (0.001*) | 0.493 (1.020) | 518 | 0.004 (0.013*) | 0.409 (1.317) | 696 | 0.032 (0.048*) | 18 | 0.206 |

*Note.* * indicates p <0.05, ** indicates p<0.001; SD = Standard Deviation; U = U statistic of the Mann-Whitney U test; $p_{FDR}$ = False Discovery Rate adjusted p-value. P-values were corrected for the different areas.

***Table S5*** Network characteristics (for 20% density) in the rest of the brain of patients with different glioma subtypes including comparison to HCs (whole brain)

| Measure, Subgroup | Delta | | | Theta | | | Lower Alpha | | |
|---|---|---|---|---|---|---|---|---|---|
| | 20% | | | 20% | | | 20% | | |
| | mean (SD) | *U* | *p* (*$p_{FDR}$*) | mean (SD) | *U* | *p* (*$p_{FDR}$*) | mean (SD) | *U* | *p* (*$p_{FDR}$*) |
| Clustering Coefficient | | | | | | | | | |
| IDH-wildtype glioblastoma | 0.470 (1.269) | 1523 | <0.001 (<0.001**) | 0.398 (1.557) | 1292 | 0.001 (0.002*) | 0.230 (1.242) | 1164 | 0.036 (0.043*) |
| IDH-mutant, 1p/19q non-codeleted | 0.330 (1.199) | 1325 | <0.001 (<0.001**) | 0.357 (1.380) | 1325 | <0.001 (<0.001**) | 0.226 (1.204) | 1135 | 0.013 (0.016*) |
| IDH-mutant, 1p/19q codeleted | 0.117 (1.176) | 635 | 0.160 (0.160) | 0.243 (1.296) | 700 | 0.028 (0.074) | 0.191 (1.250) | 650 | 0.021 (0.074) |
| HCs | 0 (0.992) | | | 0 (0.992) | | | 0 (0.992) | | |
| Eigenvector Centrality | | | | | | | | | |
| IDH-wildtype glioblastoma | -0.098 (1.111) | 504 | <0.001 (0.002*) | -0.043 (1.138) | 673 | 0.041 (0.083) | -0.011 (1.116) | 812 | 0.387 (0.464) |
| IDH-mutant, 1p/19q non-codeleted | -0.050 (1.087) | 551 | 0.008 (0.022*) | -0.023 (1.129) | 772 | 0.471 (0.707) | -0.003 (1.101) | 852 | 0.989 (0.989) |
| IDH-mutant, 1p/19q codeleted | -0.047 (1.068) | 379 | 0.093 (0.169) | -0.035 (1.089) | 483 | 0.672 (0.735) | -0.042 (1.112) | 379 | 0.093 (0.169) |
| HCs | 0 (0.992) | | | 0 (0.992) | | | 0 (0.992) | | |

*Note*. * indicates p <0.05, ** indicates p<0.001; SD = Standard Deviation; U = U statistic of the Mann-Whitney U test; *$p_{FDR}$* = False Discovery Rate adjusted p-value. P-values were corrected for the different frequency bands and densities. The means of the measures were calculated with the values standardized on the regional means and SD of HCs (dev). Therefore, for HC the mean is 0 and SD around 1.

***Table S6*** Network characteristics (for 30% density) in the rest of the brain of patients with different glioma subtypes including comparison to HCs (whole brain)

| Measure, Subgroup | Delta | | | Theta | | | Lower Alpha | | |
|---|---|---|---|---|---|---|---|---|---|
| | 30% | | | 30% | | | 30% | | |
| | mean (SD) | *U* | *p* ($p_{FDR}$) | mean (SD) | *U* | *p* ($p_{FDR}$) | mean (SD) | *U* | *p* ($p_{FDR}$) |
| Clustering Coefficient | | | | | | | | | |
| IDH-wildtype glioblastoma | 0.579 (1.282) | 1539 | <0.001 (<0.001*) | 0.431 (1.509) | 1329 | <0.001 (<0.001**) | 0.207 (1.203) | 1150 | 0.048 (0.048*) |
| IDH-mutant, 1p/19q non-codeleted | 0.372 (1.194) | 1350 | <0.001 (<0.001**) | 0.374 (1.330) | 1350 | <0.001 (<0.001**) | 0.213 (1.155) | 1102 | 0.029 (0.029*) |
| IDH-mutant, 1p/19q codeleted | 0.230 (1.195) | 709 | 0.021 (0.074) | 0.269 (1.303) | 691 | 0.037 (0.074) | 0.191 (1.179) | 655 | 0.099 (0.135) |
| HCs | 0 (0.992) | | | 0 (0.992) | | | 0 (0.992) | | |
| Eigenvector Centrality | | | | | | | | | |
| IDH-wildtype glioblastoma | -0.089 (1.135) | 481 | <0.001 (0.002*) | -0.035 (1.143) | 688 | 0.056 (0.084) | -0.003 (1.107) | 907 | 0.949 (0.949) |
| IDH-mutant, 1p/19q non-codeleted | -0.049 (1.098) | 538 | 0.005 (0.022*) | -0.017 (1.133) | 801 | 0.643 (0.771) | 0.004 (1.087) | 942 | 0.439 (0.707) |
| IDH-mutant, 1p/19q codeleted | -0.046 (1.087) | 344 | 0.035 (0.169) | -0.021 (1.090) | 490 | 0.735 (0.735) | -0.026 (1.090) | 387 | 0.113 (0.169) |
| HCs | 0 (0.992) | | | 0 (0.992) | | | 0 (0.992) | | |

*Note*. * indicates p <0.05, ** indicates p<0.001; SD = Standard Deviation; U = U statistic of the Mann-Whitney U test; $p_{FDR}$ = False Discovery Rate adjusted p-value. P-values were corrected for the different frequency bands and densities. The means of the measures were calculated with the values standardized on the regional means and SD of HCs (dev). Therefore, for HC the mean is 0 and SD around 1.

***Table S7*** Spin-test results for patients and HCs

| Measure, Group | Delta | | Theta | | Lower Alpha | |
|---|---|---|---|---|---|---|
| | 20% | 30% | 20% | 30% | 20% | 30% |
| CC and offset | | | | | | |
| Patients ($r$ [$p_{binom}$]) | 0.562 [0,<0.001]** | 0.613 [0<0.001]** | 0.598 [0, <0.001]** | 0.629 [0, <0.001]** | 0.579 [0, <0.001]** | 0.608 [0, <0.001]** |
| HCs ($r$ [$p_{binom}$]) | 0.757 [0, <0.001]** | 0.769 [0, <0.001]** | 0.705 [0, <0.001]** | 0.715 [0, <0.001]** | 0.653 [0, <0.001]** | 0.703 [0, <0.001]** |
| EC and offset | | | | | | |
| Patients ($r$ [$p_{binom}$]) | 0.124 [0.522, 0.549] | 0.099 [0.591, 0.618] | 0.405 [0.028, 0.038]* | 0.339 [0.061, 0.075] | 0.212 [0.308, 0.333] | 0.227 [0.269, 0.295] |
| HCs ($r$ [$p_{binom}$]) | 0.145 [0.402, 0.429] | 0.119 [0.481, 509] | 0.353 [0.019, 0.027]* | 0.326 [0.023, 0.033]* | 0.293 [0.192, 0.214] | 0.291 [0.190, 0.214] |

*Note*. * indicates p <0.05, ** indicates p<0.001; $r$ = Pearson's correlation; $p_{binom}$ = binomial confidence interval for the p-value.

***Table S8*** Linear Mixed Model with $offset_{dev}$ as dependent and $EC_{dev}$ and $CC_{dev}$ as independent variables including the interaction between patients and HCs to test potential differences

| Frequency, Density | Variable | Coefficient [CI] | *Z* | *p* | $p_{FDR}$ |
|---|---|---|---|---|---|
| Delta | | | | | |
| 20% | Intercept | 0 [-0,171, 0.171] | 0 | 1 | |
| | $EC_{dev}$ | 0.048 [0.032, 0.065] | 5.710 | <0.001 | <0.001** |
| | $Group_{patients}$ x $EC_{dev}$ | -0.043 [-0.064, -0.022] | -3.934 | <0.001 | <0.001** |
| | $CC_{dev}$ | 0.023 [0.005, 0.040] | 2.588 | 0.009 | 0.023* |
| | $Group_{patients}$ x $CC_{dev}$ | -0.012 [-0.033, 0.01] | -1.075 | 0.282 | 0.424 |
| 30% | Intercept | 0 [-0,171, 0.171] | 0 | 1 | |
| | $EC_{dev}$ | 0.047 [0.031, 0.063] | 5.596 | <0.001 | <0.001** |
| | $Group_{patients}$ x $EC_{dev}$ | -0.048 [-0.07, -0.027] | -4.478 | <0.001 | <0.001** |
| | $CC_{dev}$ | 0.027 [0.010, 0.044] | 3.115 | 0.002 | 0.006* |
| | $Group_{patients}$ x $CC_{dev}$ | -0.014 [-0.035, 0.007] | -1.284 | 0.199 | 0.319 |
| Theta | | | | | |
| 20% | Intercept | 0 [-0,171, 0.171] | 0 | 1 | |
| | $EC_{dev}$ | -0.008 [-0.025, 0.008] | -0.983 | 0.326 | 0.434 |
| | $Group_{patients}$ x $EC_{dev}$ | 0.022 [0, 0.043] | 1.995 | 0.046 | 0.100 |
| | $CC_{dev}$ | 0.005 [-0.012, 0.022] | 0.553 | 0.581 | 0.682 |
| | $Group_{patients}$ x $CC_{dev}$ | 0.002 [-0.019, 0.023] | 0.164 | 0.869 | 0.869 |
| 30% | Intercept | 0 [-0,171, 0.171] | 0 | 1 | |
| | $EC_{dev}$ | -0.008 [-0.025, 0.008] | -0.999 | 0.318 | 0.434 |
| | $Group_{patients}$ x $EC_{dev}$ | 0.019 [-0.002, 0.04] | 1.77 | 0.076 | 0.141 |
| | $CC_{dev}$ | 0.016 [-0.001, 0.033] | 1.795 | 0.073 | 0.141 |
| | $Group_{patients}$ x $CC_{dev}$ | -0.008 [-0.03, 0.013] | -0.745 | 0.456 | 0.576 |
| Lower Alpha | | | | | |
| 20% | Intercept | 0 [-0,171, 0.171] | 0 | 1 | |
| | $EC_{dev}$ | -0.004 [-0.020, 0.013] | -0.424 | 0.672 | 0.701 |
| | $Group_{patients}$ x $EC_{dev}$ | -0.053 [-0.074, -0.032] | -5.436 | <0.001 | <0.001** |
| | $CC_{dev}$ | 0.005[-0.013, 0.022] | 0.529 | 0.597 | 0.682 |
| | $Group_{patients}$ x $CC_{dev}$ | -0.030 [-0.051, -0.008] | -2.717 | 0.007 | 0.018* |
| 30% | Intercept | 0 [-0,171, 0.171] | 0 | 1 | |
| | $EC_{dev}$ | -0.004 [-0.020, 0.013] | -0.436 | 0.663 | 0.701 |
| | $Group_{patients}$ x $EC_{dev}$ | -0.053 [-0.074, -0.032] | -4.883 | <0.001 | <0.001** |
| | $CC_{dev}$ | 0.016 [-0,002, 0.033] | 1.719 | 0.086 | 0.147 |
| | $Group_{patients}$ x $CC_{dev}$ | -0.059 [-0.081, -0.037] | -5.321 | <0.001 | <0.001** |

*Note.* * indicates p <0.05, ** indicates p<0.001; A random intercept was fitted for participants; CI = Confidence interval for coefficient; $p_{FDR}$ = False Discovery Rate adjusted p-value. The p-values were corrected for the different frequency bands and densities. Only the independent variables were included in this correction.

***Table S9*** Linear Mixed Model with $offset_{dev}$ as dependent and $EC_{dev}$ and $CC_{dev}$ as independent variables for the whole brain of HCs

| Frequency, Density | Variable | Coefficient [CI] | *Z* | *p* | $p_{FDR}$ |
|---|---|---|---|---|---|
| Delta | | | | | |
| 20% | Intercept | 0 [-0,147, 0.147] | 0 | 1 | |
| | $EC_{dev}$ | 0.048 [0.034, 0.062] | 6.677 | <0.001 | <0.001** |
| | $CC_{dev}$ | 0.023 [0.008, 0.037] | 3.026 | 0.002 | 0.007* |
| 30% | Intercept | 0 [-0,147, 0.147] | 0 | 1 | |
| | $EC_{dev}$ | 0.047 [0.033, 0.061] | 6.543 | <0.001 | <0.001** |
| | $CC_{dev}$ | 0.027 [0.013, 0.042] | 3.642 | <0.001 | 0.001* |
| Theta | | | | | |
| 20% | Intercept | 0 [-0,147, 0.147] | 0 | 1 | |
| | $EC_{dev}$ | -0.008 [-0.023, 0.006] | -1.148 | 0.251 | 0.377 |
| | $CC_{dev}$ | 0.005 [-0.01, 0.02] | 0.645 | 0.518 | 0.622 |
| 30% | Intercept | 0 [-0,147, 0.147] | 0 | 1 | |
| | $EC_{dev}$ | -0.008 [-0.023, 0.006] | -1.166 | 0.243 | 0.377 |
| | $CC_{dev}$ | 0.016 [0.001, 0.031] | 2.096 | 0.036 | 0.087 |
| Lower Alpha | | | | | |
| 20% | Intercept | 0 [-0,147, 0.147] | 0 | 1 | |
| | $EC_{dev}$ | -0.004 [-0.018, 0.011] | -0.494 | 0.622 | 0.622 |
| | $CC_{dev}$ | 0.005 [-0.01, 0.02] | 0.616 | 0.538 | 0.622 |
| 30% | Intercept | 0 [-0,147, 0.147] | 0 | 1 | |
| | $EC_{dev}$ | -0.004 [-0.018, 0.01] | -0.508 | 0.611 | 0.622 |
| | $CC_{dev}$ | 0.016 [0, 0.031] | 2.003 | 0.045 | 0.090 |

*Note.* * indicates p <0.05, ** indicates p<0.001; A random intercept was fitted for participants; CI = Confidence interval for coefficient; $p_{FDR}$ = False Discovery Rate adjusted p-value. The p-values were corrected for the different frequency bands and densities. Only the independent variables were included in this correction.

***Table S10*** Within-subject correlation results for patients (rest of the brain) and HCs (whole brain) and their group comparison

| Measure, Group | Delta | | Theta | | Lower Alpha | |
|---|---|---|---|---|---|---|
| | 20% | 30% | 20% | 30% | 20% | 30% |
| $CC_{dev}$ and $offset_{dev}$ | | | | | | |
| Patients rest of the brain ($r$ ($p$, $p_{FDR}$)) | 0.032 (0.789, 0.789) | 0.006 (0.721, 0.789) | 0.022 (0.099, 0.179) | 0.019 (0.119, 0.179) | -0.035 (0.043, 0.130) | -0.049 (0.007, 0.046*) |
| HCs ($r$ ($p$, $p_{FDR}$)) | 0.029 (0.014, 0.037*) | 0.030 (0.019, 0.037*) | 0.014 (0.195, 0.233) | 0.028 (0.017, 0.037*) | 0.008 (0.419, 0.419) | 0.018 (0.144, 0.216) |
| Group comparison ($U$ ($p$, $p_{FDR}$)) | 2222 (0.174, 0.280) | 2232 (0.187, 0.280) | 2671 (0.664, 0.681) | 2459 (0.681, 0.681) | 2072 (0.049, 0.149) | 1818 (0.003, 0.017*) |
| $EC_{dev}$ and $offset_{dev}$ | | | | | | |
| Patients rest of the brain ($r$ ($p$, $p_{FDR}$)) | 0.002 (0.643, 0.643) | -0.004 (0.489, 0.587) | 0.021 (0.225, 0.397) | 0.017 (0.265, 0.397) | -0.065 (0.009, 0.041*) | -0.058 (0.014, 0.041*) |
| HCs ($r$ ($p$, $p_{FDR}$)) | 0.066 (<0.001, <0.001**) | 0.063 (<0.001, <0.001**) | 0.001 (0.653, 0.986) | -0.001 (0.771, 0.986) | -0.007 (0.986, 0.986) | -0.008 (0.928, 0.986) |
| Group comparison ($U$ ($p$, $p_{FDR}$)) | 1717 (<0.001, 0.004*) | 1756 (0.001, 0.003*) | 2685 (0.624, 0.624) | 2714 (0.544, 0.624) | 2113 (0.072, 0.124) | 2128 (0.082, 0.124) |

*Notes.* * indicates p <0.05, ** indicates p<0.001; r = Pearson's correlation; $p_{FDR}$ = False Discovery Rate adjusted p-value. The p-values were corrected for the different frequency bands and densities.

***Table S11*** Linear Mixed Model with $offset_{dev}$ as dependent variable and lower alpha $EC_{dev}$ and $CC_{dev}$ as independent variables for the different subtypes of glioma (rest of the brain)

| Frequency, Density | Variable | Coefficient [CI] | Standardized Coefficient (betas) | *Z* | *p* | $p_{FDR}$ |
|---|---|---|---|---|---|---|
| IDH-wt glioblastoma | | | | | | |
| Lower Alpha | | | | | | |
| 20% | Intercept | 0.349 [0.063, 0.635] | | 2.393 | 0.017 | |
| | $EC_{dev}$ | -0.118 [-0.144, -0.092] | -0.098 | -8.812 | <0.001 | <0.001** |
| | $CC_{dev}$ | -0.021 [-0.046, 0.005] | -0.019 | -1.579 | 0.114 | 0.114 |
| 30% | Intercept | 0.357 [0.072, 0.642] | | 2.452 | 0.014 | |
| | $EC_{dev}$ | -0.101 [-0.127, -0.075] | -0.083 | -7.646 | <0.001 | <0.001** |
| | $CC_{dev}$ | -0.058 [-0.084, -0.032] | -0.051 | -4.363 | <0.001 | <0.001** |
| IDH-mutant, 1p19q non-codeleted | | | | | | |
| Lower Alpha | | | | | | |
| 20% | Intercept | 0.402 [0.135, 0.669] | | 2.954 | 0.003 | |
| | ECdev | 0.008 [-0.016, 0.031] | 0.007 | 0.659 | 0.509 | 0.547 |
| | CCdev | 0.007 [-0.16, 0.03] | 0.007 | 0.602 | 0.547 | 0.547 |
| 30% | Intercept | 0.136 [0.134, 0.668] | | 2.947 | 0.003 | |
| | ECdev | 0.017 [-0.007, 0.04] | 0.015 | 1.38 | 0.167 | 0.547 |
| | CCdev | 0.012 [-0.012, 0.036] | 0.012 | 0.951 | 0.342 | 0.547 |
| IDH-mutant, 1p19q codeleted | | | | | | |
| Lower Alpha | | | | | | |
| 20% | Intercept | 0.452 [0.071, 0.834] | | 2.325 | 0.020 | |
| | $EC_{dev}$ | -0.066 [-0.102, -0.031] | -0.056 | -3.647 | <0.001 | <0.001** |
| | $CC_{dev}$ | -0.018 [-0.051, 0.015] | -0.017 | -1.069 | 0.285 | 0.285 |
| 30% | Intercept | 0.456 [0.076, 0.837] | | 2351 | 0.019 | |
| | $EC_{dev}$ | -0.069 [-0.105, -0.034] | -0.057 | -3.838 | <0.001 | <0.001** |
| | $CC_{dev}$ | -0.035 [-0.07, -0.001] | -0.032 | -1.991 | 0.047 | 0.062 |

*Note.* * indicates p <0.05, ** indicates p<0.001; A random intercept was fitted for participants; CI = Confidence interval for coefficient; $p_{FDR}$ = False Discovery Rate adjusted p-value. The p-values were corrected for the two densities. Only the independent variables were included in this correction.

***Table S12*** Within-subject correlation results for the different glioma subgroups (for the lower Alpha band)

| Subtype | $CC_{dev}$ and $offset_{dev}$ | | $EC_{dev}$ and $offset_{dev}$ | |
|---|---|---|---|---|
| | 20% | 30% | 20% | 30% |
| IDH-wildtype glioblastoma | -0.0345 | -0.061 | -0.100 | -0.090 |
| ($r$ ($p$, $p_{FDR}$)) | (0.382, 0.382) | (0.100, 0.200) | (0.019, 0.029*) | (0.029, 0.029*) |
| IDH-mutant, 1p/19q non-codeleted | 0.012 | 0.010 | 0.012 | 0.021 |
| ($r$ ($p$, $p_{FDR}$)) | (0.779, 0.849) | (0.849, 0.849) | (0.831, 0.831) | (0.582, 0.831) |
| IDH-mutant, 1p/19q co-deleted | -0.055 | -0.070 | -0.101 | -0.098 |
| ($r$ ($p$, $p_{FDR}$)) | (0.159, 0.159) | (0.079, 0.159) | (0.020, 0.020*) | (0.008, 0,016*) |

*Note.* * indicates p <0.05, ** indicates p<0.001; r = Pearson's correlation; $p_{FDR}$ = False Discovery Rate adjusted p-value. The p-values were corrected for the two densities.

***Table S13*** Correlations between peritumoral offset and associations in the rest of the brain

| Subtype | Peritumoral Area all regions | | | | Peritumoral Area 3 highest activity regions | | | |
|---|---|---|---|---|---|---|---|---|
| | $CC_{dev}$ and $offset_{dev}$ Lower Alpha | | $EC_{dev}$ and $offset_{dev}$ Lower Alpha | | $CC_{dev}$ and $offset_{dev}$ Lower Alpha | | $EC_{dev}$ and $offset_{dev}$ Lower Alpha | |
| | 20% | 30% | 20% | 30% | 20% | 30% | 20% | 30% |
| Patients whole group (*r (p, $p_{FDR}$))* | -0.027 (0.829, 0.829) | -0.065 (0.601, 0.829) | -0.058 (0.638, 0.638) | -0.076 (0.542, 0.638) | 0.003 (0.979, 0.979) | -0.008 (0.947, 0.979) | 0.029 (0.813, 0.874) | 0.019 (0.874, 0.874) |
| IDH-wildtype glioblastoma (*r* (*p*, $p_{FDR}$)) | 0.063 (0.769, 0.944) | 0.015 (0.943, 0.943) | -0.222 (0.296, 0.338) | -0.204 (0.338, 0.338) | 0.129 (0.545, 0.579) | 0.119 (0.579, 0.579) | -0.134 (0.532, 0.606) | -0.111 (0.606, 0.606) |
| IDH-mutant, 1p/19q non-codeleted (*r* (*p*, $p_{FDR}$)) | -0.203 (0.353, 0.353) | -0.209 (0.339, 0.353) | -0.055 (0.803, 0.803) | -0.108 (0.622, 0.803) | -0.198 (0.366, 0.366) | -0.199 (0.361, 0.366) | -0.059 (0.787, 0.682) | -0.090 (0.682, 0.787) |
| IDH-mutant, 1p/19q-codeleted (*r* (*p*, $p_{FDR}$)) | -0.003 (0.993, 0.993) | -0.182 (0.591, 0.993) | 0.138 (0.685, 0.692) | 0.135 (0.692, 0.692) | -0.123 (0.717, 0.717) | -0.303 (0.363, 0.717) | 0.145 (0.670, 0.739) | 0.114 (0.739, 0.739) |

*Notes.* * indicates p <0.05, ** indicates p<0.001; r = Pearson's correlation; $p_{FDR}$ = False Discovery Rate adjusted p-value. The p-values were corrected for the two densities.
